# GENDER NORMS AND ATTITUDE TOWARDS ABORTION AMONG VERY YOUNG ADOLESCENTS IN KENYA AND NIGERIA

**DOI:** 10.1101/2024.04.08.24305479

**Authors:** Matthew Ayodele Alabi, Bamidele M. Bello, Beatrice W. Maina

**Affiliations:** Population Council, Abuja, Nigeria; Department of Public Health, University of Ibadan, Nigeria; African Population and Health Centre, Nairobi, Kenya

**Keywords:** gender norms, Very young adolescents, abortion, attitude

## Abstract

**Introduction:** Unsafe abortion is a major cause of death in sub-Saharan African countries with very young adolescents (VYAs) at increased risk due to their high vulnerability to unprotected sex and unplanned pregnancies. Abortion beliefs and attitudes are considered to be partly rooted in traditional views on gender and religious influences. This study is informed by the limited data on gender norm perception and its association with abortion among VYAs despite the increasing prevalence of unsafe abortion reported among this group.

**Materials:** Data for this study was collected as part of a longitudinal survey on the gendered socialization and sexual and reproductive health of very young, in-school adolescents aged 10-14 years in Kenya and Nigeria. The study obtained quantitative data from 1,912 VYAs using a structured questionnaire. The results presented in this paper are from the quantitative baseline data collected in Kenya and Nigeria

**Result:** The study found significant regional differentials in attitudes toward abortion and gender norm perception of the VYAs from the two regions. VYAs from Nigeria were more likely to endorse abortion practices relative to their counterparts from Kenya. Factors associated with endorsement of abortion practice were gender norms about Sexual Double Standards (SDS) and Normative Heterosexual Relation (NHR) in Nigeria and knowledge of where to get a condom, NHR, and Normative Romantic Relationship (NRR) in Kenya.

**Conclusion:** Intervention efforts seeking to promote positive gender norms and attitudes towards SRH must begin with the VYAs and must consider regional variations and address knowledge and access to SRH commodities.

**What is already known on this topic:** previous studies have explored attitudes towards gender norms and abortion among young people and adults, with little known among very young adolescents aged 10-14 years.

**What this study adds:** the study contributed to the limited research on gender norms and attitudes towards abortion among very young adolescents.

**How this study might affect research, practice, and policy:** The outcome of this suggests the need to consider regional variations when developing interventions and policies addressing gender norms and attitudes towards abortion in Africa.

## Background

About 73 million induced abortions take place globally annually (1). Approximately 45% of all induced abortions are unsafe, with developing countries bearing the highest burden of unsafe abortion (95%), more so in Africa and Latin America (1). Unsafe abortion according to the World Health Organization refers to those performed by individuals either lacking the requisite skills or in an environment that does not follow minimal medical standards or both, in addition to highlighting how social and legal contexts influence abortion safety (2). Abortion among adolescent girls constitutes a considerable proportion of the abortion rate globally, with 15% of unsafe abortions occurring among adolescent girls under age 20 years (3). Nearly three-quarters of abortion-induced hospitalization reported in developing countries are among adolescent girls under age 20 years (4). Although abortion is considered illegal in several African countries including Kenya and Nigeria, except on certain stringent conditions such as life-threatening cases, opposition to abortion is considered to be partly rooted in traditional views on gender and religious influences (5,6).

In Kenya and Nigeria, the attitude towards abortion is generally conservative. While the constitutions of both countries suggest a possibility of women accessing safe abortion(7), there remains a negative sociocultural attitude towards abortion in these countries (8). The negative attitude towards abortion no doubt can generate undesirable sexual and reproductive health (SRH) outcomes. For instance, a Kenyan study (8) found a varied and high prevalence of negative attitudes toward abortion influenced by several socioeconomic and cultural variables including gender, place of residence, level of education and position in society among others. Similarly, a study examining attitudes and practices of abortion among female in-school adolescents in Lagos, Southwest, Nigeria found over eighty percent of them had knowledge of abortion and nearly all of them (99.2%) disapproved of abortion practice. The study found that while knowledge of abortion legality and procedure was high among the very young adolescents (age 10 – 14) (VYA), they were less likely to approve of the practice relative to their older counterparts in the age group 15-19 years (9).

Gender norms - socially constructed roles, behaviour, activities, and attributes considered appropriate for boys/men or girls/women in a particular society (American Psychological Association, 2011) - are usually acquired during the early stages of childhood development (especially during the early adolescent period) through social interactions, within a particular setting. The adolescence period is generally seen as a very critical period in an individual’s life largely because identities are formed during this stage of life and a number of attitudes and behaviour become noticeable (10). Scholars have argued that gender norms can be acquired at a much earlier childhood stage of development. For instance, it is believed that by age six, children already possess the ability to distinguish between what it means to be a boy or a girl, and the attributes of each sex (11,12). This is in addition to the ability to express appropriate sex-role preferences for themselves and behave in accordance with sex-role standards (13).

Previous studies in America, Asia and Africa have reported a significant positive association between gender norm perception and attitude towards abortion (14,15). In a South African study, the authors were of the view that attitude towards abortion was clearly linked with socioeconomic and gender ideological dynamics and social status. Besides, not until recently, most studies on abortion (17,18) have particularly focused on young women aged 15 to 24 years and older adults, while neglecting the VYA. Although prevalence of sexual activity may not be as high among VYA, early adolescence is a stage where sexual activity is initiated and beliefs and norms about sexual relations and activities internalized. (19–21). Findings from the literature suggest gender norms and power imbalance relating to women’s SRH are largely influenced by the absence of agency in making SRH decisions, stigma, lack of financial resources and the need to be secretive often results in harmful abortion practices and also delays in seeking care (22).

However, to the best of our knowledge, studies linking gender norms with attitude towards abortion in Africa and especially among VYA remain very sparse. Hence, given the high rate of risky sexual activity among older adolescents and the pathways though which norms and attitudes are developed, determining the attitudes towards abortion and the association these attitudes gender norms and SRH behaviour of these VYA will no doubt help in developing effective programmatic responses aimed at addressing the SRH needs of VYA.

## MATERIAL AND METHODS

### Study Design/Population

The study utilized baseline data from a longitudinal survey on the gendered socialization of very young in-school adolescents and their sexual and reproductive health, conducted in urban informal settlements in Nigeria (Ile-Ife and Modakeke), and Kenya (Korogocho and Viwandani). The survey adopted a concurrent mixed-method research design, which involves the collection of both quantitative and qualitative data. The data collection tools were developed and validated in the context of the <u>Global Early Adolescent Study</u> and comprises of three instruments: a health+ instrument, a vignette-based instrument, and the gender norms scale. The tool has undergone face and content validity assessment and has been pilot tested in a previous GEAS study in Ile-Ife, Nigeria and Korogocho slums, Kenya (23).

### Data Collection

For this study, we used the health+ instrument and the gender norm scale to measure gender norms and SRH outcomes among VYA. The health+ instrument obtained socio-demographic information and a number of key health and behavioral factors including pubertal maturation; romantic relationships; mental health and future expectation. Measures of abortion and the socio-demographic characteristics of the early adolescents were obtained from this instrument. The gender-norm scale assessed the gendered perception of adolescents using a series of questions that have gender undertone. Information collected through the use of this instruments were used to assess early adolescents’ gender-related beliefs. Data were collected using computer-assisted personal interviews (CAPI) on tablets and uploaded to a central data repository, where all data files were maintained in a secure server. Identifiers were removed from all analytical datasets and from the presentation of results.

### Sampling Technique

The study was conducted in seven randomly selected schools serving primarily urban disadvantaged population within Ife and Modakeke community in Ile-Ife, an ancient city in Osun State, southwest Nigeria and four schools in Korogocho and Viwandani informal settlements in Kenya. A total of 1,032 and 880 in-school VYA in Nigeria and Kenya were recruited and interviewed.

### Measurement of Variables

#### Outcome Variable

The outcome variable for this study is attitude towards abortion measured using the perception of the adolescents towards abortion. Three questions were asked namely: Adolescent girls who get pregnant should have an abortion if they are not married; Adolescent girls who get pregnant should have an abortion because they are too young to raise children; Adolescent girls who get pregnant should have an abortion to stay in school. All the three questions were measured on a five-point Likert scale, ranging from disagree a lot coded as 1 and agree a lot coded as 5. However, for ease of analysis principal component analysis (PCA) was performed and used to generate a single score for all the three items and grouped into high and low. High scores indicate acceptance of abortion (and coded as 1), while low score indicates disapproval for abortion (coded as 0).

#### Explanatory Variables

The principal explanatory variable in this study is gender norms. The overall gender norm aspect of the multidimensional scale comprises of three sub-scales namely: sexual double standard scale, stereotypical view about toughness versus weakness and normative heterosexual relation. The sexual double standard scale was further divided into three domains namely: sexual double standard (SDS), normative romantic relation (NRR) and masculine sexual prowess (MSP). Each of the scales were measured on a 5-point Likert scale ranging from disagree a lot (1), disagree a little (2), neither agree nor disagree (3), agree a little (4) to agree a lot (5). The reliability analysis of the scales yielded a Cronbach alpha exceeding the minimum recommended value of 0.70 except for MSP sub-domain: SDS scale (α=0.89), stereotypical view about toughness versus weakness (α=0.82); normative heterosexual relation scale (α=0.85); SDS sub scale (α=0.86); NRR sub-scale (α=0.78); MSP (α=0.53) and attitude towards abortion (α=0.89). Since the MSP subscale failed to reach the minimum acceptable threshold, it was excluded from further analysis. Other explanatory variables used include socio-demographic characteristics namely: age, religion (Christian versus Islam) and sex (male versus female), knowledge of where to get condom and pregnancy prevention methods, with yes or no as the options.

### Ethical consideration

To ensure the research conforms to the highest scientific and ethical standards, the research protocol was submitted for review and approved by local ethics boards of the respective countries. Given the nature of the information to be gathered, protecting, and respecting the confidentiality of our informants was a critical consideration throughout the study. Therefore, all participants were adequately informed about the purpose of the study and methods to be used; anticipated indirect benefits, the lack of direct benefits such as material compensation; potential risks; the right to abstain from participating in the study, or to withdraw from it at any time, without reprisal. Both informed parental consent and adolescent assent were obtained for each participant prior to interviewing adolescents.

### Statistical Analysis

Both descriptive and inferential analysis were performed. Univariate analysis involved performing frequency count and percentages. Bivariate analysis involved performing Chi square test and test of difference (independent t-test and one-way ANOVA), while multivariate analysis involved performing Principal Component Analysis and Binary Logistic Regression. For the multivariate binary logistic regression, the odds ratio was reported, while the goodness of fit of the model was used to determine whether the application of binary logistic regression followed the required assumption and whether the model sufficiently fits the data. Only explanatory variables that were statistically significant with the outcome variable at the bivariate level were included in the multivariate model. The Hosmer-Lemeshow test which checks for goodness of fit for the logistic regression model was used, with small Chi-square and large p-value indicating a good fit to the data (24). For our study, the Hosmer-Lemeshow statistics for the combined model was (χ^2^=296.91, p=0.635), the Nigeria model (χ^2^=139.24, p=0.25), and Kenya model (χ^2^=232.55, p=0.78). Hence, our goodness of fit test based on the p-value obtained indicates the model adequately fits the data.

## RESULTS

### Descriptive Statistics

Table 1 presents the sociodemographic characteristics of the study participants. The mean age of the respondents was 12.0, SD=1.2. (Nigeria 12.0, SD=1.3; Kenya 12.0, SD=1.1). Respondents who are 12 years of age accounted for a higher proportion. While the study participants in Nigeria were evenly distributed by sex, a higher proportion of the participants in Kenya were females (58%).

**Table 1:**
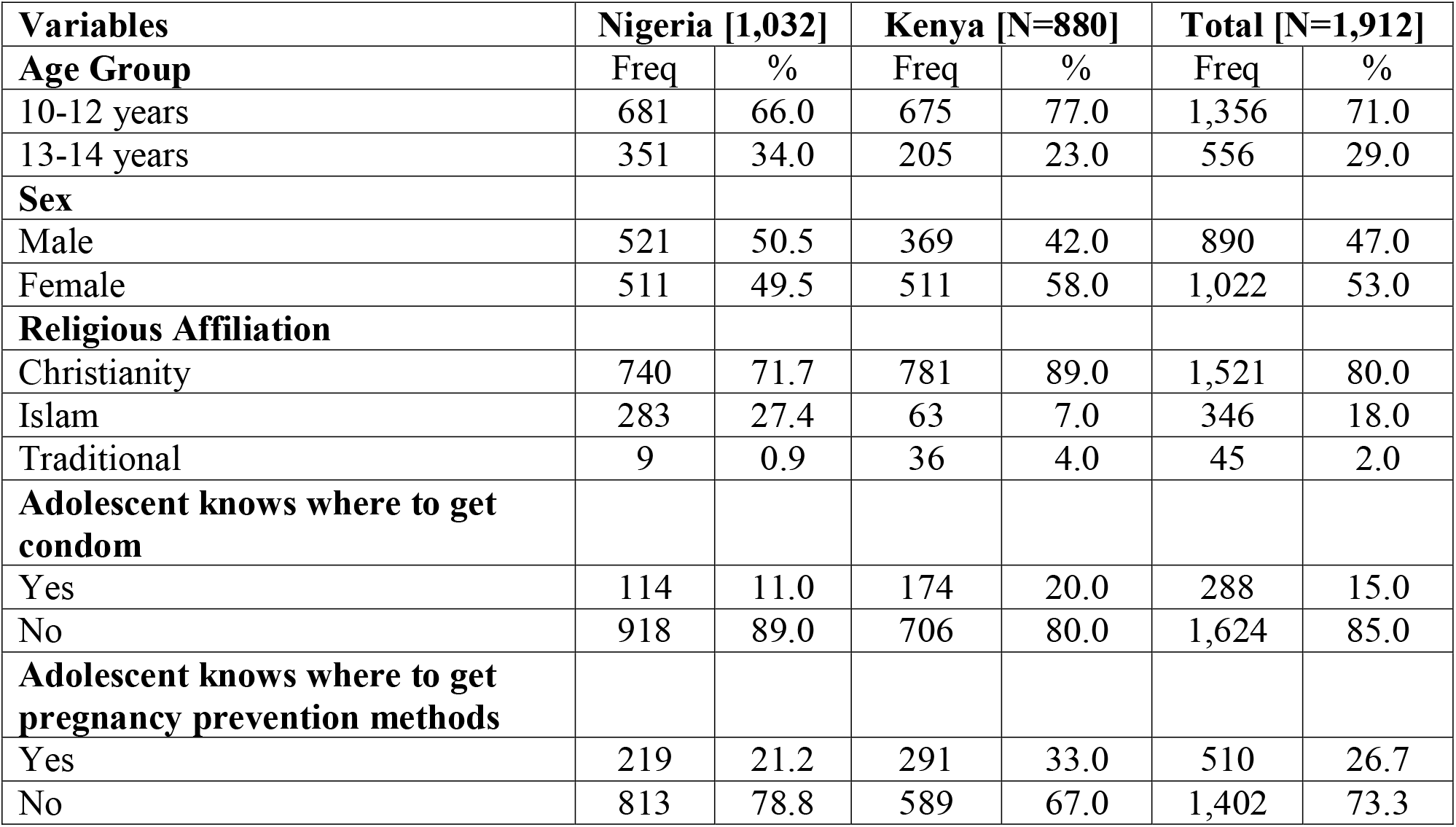
Socio-demographic and SRH outcome.

| **Variables** | **Nigeria [1,032]** |  | **Kenya [N=880]** |  | **Total [N=1,912]** |  |
| --- | --- | --- | --- | --- | --- | --- |
| **Age Group** | Freq | % | Freq | % | Freq | % |
| 10-12 years | 681 | 66.0 | 675 | 77.0 | 1,356 | 71.0 |
| 13-14 years | 351 | 34.0 | 205 | 23.0 | 556 | 29.0 |
| **Sex** |  |  |  |  |  |  |
| Male | 521 | 50.5 | 369 | 42.0 | 890 | 47.0 |
| Female | 511 | 49.5 | 511 | 58.0 | 1,022 | 53.0 |
| **Religious Affiliation** |  |  |  |  |  |  |
| Christianity | 740 | 71.7 | 781 | 89.0 | 1,521 | 80.0 |
| Islam | 283 | 27.4 | 63 | 7.0 | 346 | 18.0 |
| Traditional | 9 | 0.9 | 36 | 4.0 | 45 | 2.0 |
| **Adolescent knows where to get condom** |  |  |  |  |  |  |
| Yes | 114 | 11.0 | 174 | 20.0 | 288 | 15.0 |
| No | 918 | 89.0 | 706 | 80.0 | 1,624 | 85.0 |
| **Adolescent knows where to get pregnancy prevention methods** |  |  |  |  |  |  |
| Yes | 219 | 21.2 | 291 | 33.0 | 510 | 26.7 |
| No | 813 | 78.8 | 589 | 67.0 | 1,402 | 73.3 |

### Gender Norm Perception

Table 2 presents gender norm perception among the VYAs from Kenya and Nigeria. The result of the Chi-Square test revealed significant differences in gender norm perception between the VYA from the Kenya and Nigeria (p<0.05) for all the dimensions and sub-domains of gender norms examined. Regional variation suggests a stronger endorsement of these gender norms among VYA in the Nigeria relative to their Kenyan counterparts. For instance, with respect to Sexual Double Standard (SDS) scale, there were more VYA from Nigeria who scored high relative to their Kenyan counterpart. Similarly, there were more VYA from Nigeria who scored high on stereotypical views about toughness versus weakness, Normative Heterosexual Relation, and Normative Romantic Relation relative to their Kenyan counterpart.

**Table 2:** Gender Norm Perception and Attitude towards abortion.

| **Gender norm scales** | **Nigeria [1,032]** |  | **Kenya [N=880]** |  | **Total [N=1,912]** |  |
| --- | --- | --- | --- | --- | --- | --- |
| **Sexual Double Standard (SDS) Scale** | **Freq** | **%** | **Freq** | **%** | **Freq** | **%** |
| Low | 225 | 21.8 | 425 | 48.3 | 650 | 34.0 |
| Moderate | 323 | 31.2 | 325 | 37.0 | 648 | 33.9 |
| High | 484 | 47.0 | 130 | 14.7 | 614 | 32.1 |
| | $\chi^2=225.17, p=0.001$ | | | | | |
| **Stereotypical View About Toughness Versus Weakness Scale** |  |  |  |  |  |  |
| Low | 150 | 14.6 | 506 | 57.5 | 656 | 34.3 |
| Moderate | 472 | 45.7 | 266 | 30.2 | 738 | 38.6 |
| High | 410 | 39.7 | 108 | 12.3 | 518 | 27.1 |
| | $\chi^2=417.32, p=0.001$ | | | | | |
| **Normative Heterosexual Relation (NHR) Scales** |  |  |  |  |  |  |
| Low | 274 | 26.6 | 388 | 44.1 | 662 | 34.6 |
| Moderate | 344 | 33.3 | 284 | 32.3 | 628 | 32.9 |
| High | 414 | 40.1 | 208 | 23.6 | 622 | 32.5 |
| | $\chi^2=82.02, p=0.001$ | | | | | |
| **Normative Romantic Relation (NRR) sub scale** |  |  |  |  |  |  |
| Low | 328 | 31.8 | 388 | 44.0 | 716 | 37.4 |
| Moderate | 270 | 26.1 | 320 | 36.4 | 590 | 30.9 |
| High | 434 | 42.1 | 172 | 19.6 | 606 | 31.7 |
| | $\chi^2=111.16, p=0.001$ | | | | | |
| **Attitude towards abortion** |  |  |  |  |  |  |
| **Adolescent girls who get pregnant should have an abortion if they are not married** | **Freq** | **%** | **Freq** | **%** | **Freq** | **%** |
| Agree | 434 | 42.0 | 349 | 40.0 | 783 | 41.0 |
| Neutral | 30 | 3.0 | 87 | 10.0 | 117 | 6.0 |
| Disagree | 568 | 55.0 | 444 | 50.0 | 1,012 | 53.0 |
| | $\chi^2=40.36, p=0.001$ | | | | | |
| **Adolescent girls who get pregnant should have an abortion because they are too young to raise children** |  |  |  |  |  |  |
| Agree | 500 | 48.4 | 396 | 45.0 | 896 | 46.9 |
| Neutral | 30 | 3.0 | 85 | 10.0 | 115 | 6.0 |
| Disagree | 502 | 48.6 | 399 | 45.0 | 901 | 47.1 |
| | $\chi^2=38.31, p=0.001$ | | | | | |
| **Adolescent girls who get pregnant should have an abortion to stay in school** |  |  |  |  |  |  |
| Agree | 510 | 49.4 | 430 | 48.9 | 940 | 49.2 |
| Neutral | 36 | 3.5 | 91 | 10.3 | 127 | 6.6 |
| Disagree | 486 | 47.1 | 359 | 40.8 | 845 | 44.2 |
| | $\chi^2=37.87, p=0.001$ | | | | | |

### Attitude towards Abortion

The attitude of the VYAs towards abortion was statistically significantly different for all the measures between Kenya and Nigeria (Table 3). For instance, 42% and 40%, of VYAs from Nigeria and Kenya respectively agreed that adolescent girls who get pregnant should have an abortion if they are not married. Similarly, slightly higher proportion of VYA from Nigeria than in Nairobi were in support that adolescent girls who get pregnant should have an abortion because they are too young to raise children. However, there was a statistically significant difference in the proportion of VYA from Nigeria and Kenya who agreed that adolescent girls who get pregnant should have an abortion to stay in school.

**Table 3:** Gender norms and Abortion Perception among VYA.

| Gender Norms | Nigeria |  |  | Kenya |  |  |
| --- | --- | --- | --- | --- | --- | --- |
|  | Attitude towards abortion |  |  |  |  |  |
| *Sexual Double Standard Scale (SDS)* | Disapproval | Neutral | Approval | Disapproval | Neutral | Approval |
| Low | 33.0 | 20.0 | 10.0 | 54.0 | 42.0 | 52.0 |
| Moderate | 27.0 | 34.0 | 34.0 | 32.0 | 42.0 | 34.0 |
| High | 40.0 | 46.0 | 56.0 | 14.0 | 16.0 | 14.0 |
| | $\chi^2=57.66, p=0.001$ | | | $\chi^2=11.16, p=0.025$ | | |
| *Stereotypical View About Toughness Versus Weakness Scale* |  |  |  |  |  |  |
| Low | 17.0 | 16.0 | 10.0 | 56.0 | 56.0 | 62.0 |
| Moderate | 43.0 | 49.0 | 46.0 | 32.0 | 31.0 | 28.0 |
| High | 40.0 | 35.0 | 44.0 | 12.0 | 13.0 | 10.0 |
| | $\chi^2=11.21, p=0.024$ | | | $\chi^2=2.73, p=0.605$ | | |
| *Normative Heterosexual Relation Scales* |  |  |  |  |  |  |
| Low | 39.0 | 23.0 | 16.0 | 49.0 | 38.0 | 48.0 |
| Moderate | 34.0 | 35.0 | 31.0 | 29.0 | 36.0 | 30.0 |
| High | 27.0 | 42.0 | 53.0 | 22.0 | 26.0 | 22.0 |
| | $\chi^2=70.00, p=0.001$ | | | $\chi^2=10.68, p=0.030$ | | |
| Sub domains |  |  |  |  |  |  |

| **Normative Romantic Relation** |  |  |  |  |  |  |
| --- | --- | --- | --- | --- | --- | --- |
| Low | 41.5 | 28.2 | 24.1 | 55.0 | 39.0 | 40.0 |
| Moderate | 25.4 | 29.0 | 24.6 | 31.0 | 39.0 | 38.0 |
| High | 33.1 | 42.8 | 51.3 | 14.0 | 22.0 | 22.0 |
| | $\chi^2=34.69, p=0.001$ | | | $\chi^2=16.98, p=0.002$ | | |

### Gender norm perception and attitude towards Abortion Practice

Table 3 assesses the association between gender norm perception and attitude towards abortion among the VYA. The result revealed a statistically significant association (p<0.05) between all the dimensions and sub-domains of gender norms and abortion perception. More VYA who scored high on the SDS scale, stereotypical view about toughness versus weakness, NRR, and MSP from Nigeria were more likely to be in support of abortion relative to their counterpart from Kenya.

### Multivariate Analysis

The result of the multivariate analysis is presented in Table 4. The combined model revealed knowledge of where to get condoms and NRR were the only explanatory variables that significantly predicted attitudes toward abortion. The VYAs who knew where to get a condom demonstrated higher odds (OR=1.36, 95% C.I. 0.99-1.85) of endorsing abortion among adolescent girls. Also, VYA who scored between moderate (OR=1.49, 95% C.I. 1.06-2.06) and high (OR=1.97, 95% C.I. 1.26-3.01) on NRR were more likely to endorse abortion relative to those who scored low.

**Table 4:** Binary logistic regression showing predictors of attitude towards abortion.

| **Explanatory variables** | **Nigeria** |  | **Kenya** |  | **Total** |  |
| --- | --- | --- | --- | --- | --- | --- |
|  | **OR** | **95% CI** | **OR** | **95% CI** | **OR** | **95% CI** |
| **Sex** |  |  |  |  |  |  |
| Male | 1.000 |  | 1.000 |  | 1.000 |  |
| Female | 1.081 | 0.66-1.76 | 0.876 | 0.64-1.20 | 0.918 | 0.71-1.19 |
| **Adolescent knows where to get condom** |  |  |  |  |  |  |
| No | 1.000 |  |  |  | 1.000 |  |
| Yes | 1.073 | 0.62-1.85 | 1.533* | 1.04-2.25 | 1.360* | 0.99-1.85 |
| **Adolescents knows where to get pregnancy prevention methods** |  |  |  |  |  |  |
| No | 1.000 |  | 1.000 |  | 1.000 |  |
| Yes | 1.256 | 0.72-1.18 | 1.164 | 0.83-1.63 | 1.174 | 0.89-1.55 |
| **Sexual Double Standard (SDS)** |  |  |  |  |  |  |
| **Scale** |  |  |  |  |  |  |
| Low | 1.000 |  | 1.000 |  | 1.000 |  |
| Moderate | 3.418** | 1.25-9.31 | 0.923 | 0.62-1.37 | 1.137 | 0.81-1.61 |
| High | 1.864 | 0.60-5.81 | 0.567 | 0.30-1.04 | 0.748 | 0.46-1.22 |
| **Stereotypical View About Toughness Versus Weakness Scale** |  |  |  |  |  |  |
| Low | 1.000 |  | 1.000 |  | 1.000 |  |
| Moderate | 0.778 | 0.33-1.82 | 1.108 | 0.77-1.59 | 0.943 | 0.69-1.28 |
| High | 1.463 | 0.61-3.49 | 1.067 | 0.64-1.77 | 1.281 | 0.88-1.87 |
| **Normative Heterosexual Relation Scales** |  |  |  |  |  |  |
| Low | 1.000 |  | 1.000 |  |  |  |
| Moderate | 1.491 | 0.67-3.08 | 0.621* | 0.43-0.90 | 0.740 | 0.53-1.02 |
| High | 2.581* | 1.14-5.86 | 0.840 | 0.54-1.31 | 1.104 | 0.76-1.60 |
| **Normative Romantic Relationship** |  |  |  |  |  |  |
| Low | 1.000 |  | 1.000 |  | 1.000 |  |
| Moderate | 0.939 | 0.42-2.08 | 1.66** | 1.13-2.46 | 1.491* | 1.06-2.06 |
| High | 1.034 | 0.45-2.37 | 2.48** | 1.41-4.38 | 1.967** | 1.26-3.01 |

\*Statistically significant at 5%, \*\* statistically significant at 1%

Furthermore, factors predicting attitudes towards abortion were found to differ according to region. Among VYAs from Nigeria, SDS and NHR were the two gender norm behaviour that significantly predicted attitudes towards abortion. The VYAs from Nigeria with moderate score on SDS were about four times (OR=3.42, 95% C.I. 1.25-9.31) more likely to endorse abortion among adolescent girls relative to those who scored low. Similarly, VYAs who scored high on NHR were about three times (OR=2.58, 95% C.I. 1.14-5.86) more likely to endorse abortion practice when compared to those who scored low.

Among the VYAs from Kenya, knowledge of where to get condoms, NHR and NRR significantly predicted attitudes towards abortion. VYAs who knew where to get condoms were 1.53 times (OR=1.53, 95% C.I. 1.04-2.25) more likely to endorse abortion among adolescent girls. On the other hand, while moderate scores on NHR were associated with lower odds of endorsing abortion (OR=0.62, 95% C.I. 0.43-0.90) relative to those who scored low, a high score on NRR was associated with higher odds of endorsing abortion. VYAs who scored between moderate (OR=1.66, 95% C.I. 1.13-2.46) to high score (OR=2.48, 95% C.I. 1.41-4.38) on NRR were more likely to endorse abortion relative to those who scored low.

## Discussion

Our study found significant differences in gender norm perceptions and attitudes towards abortion for both countries and these gender norms were significantly associated with attitudes towards abortion. Adolescents, like adults, are influenced by cultural and societal expectations around gender roles and behaviors (25). These norms can shape their beliefs and attitudes on various issues (26), including abortion. Traditional gender norms may lead to different attitudes towards abortion among adolescent boys and girls. Adolescent boys raised from cultures that emphasize traditional masculinity may be socialized to adopt more conservative views on issues related to sexuality and reproduction, including abortion. They may be encouraged to prioritize notions of responsibility and control, which can lead to more restrictive attitudes towards abortion (26). On the other hand, adolescent girls may face expectations related to motherhood and family planning. These expectations could influence their attitudes towards abortion, with some girls feeling pressured to conform to traditional roles and views on reproductive choices. For example, (27) observed that despite the decriminalisation of abortion in Mozambique, young women’s lack of autonomy to make a decision on pregnancy termination was still a significant barrier to accessing safe abortion services.

Attitudes towards abortion are complex and can vary widely among adolescents. Not all adolescents conform to traditional gender norms, and individual experiences, education, exposure to different life perspectives, and personal beliefs also play significant roles in shaping their attitudes towards abortion. For instance, in a study on stigma and social norms in women’s abortion experiences among young women in Kenya and India^i^, young women demonstrated divergent views with respect to abortion. While a few of the participants held positive views for example stating that a woman should be able to decide what happens to her body, majority of the participants expressed their gendered opinion towards abortion considering women who seek abortion as irresponsible” or “immoral. Opinions such as the latter are often born from religious views that prohibit premarital sex and/or abortion or from a gendered lens promoting sexual double standards that normalises premarital sex for males as against females (28,29).

Our study found less acceptance of abortion practice among adolescents in both Nigeria and Kenya. Previous studies (9,30) in both countries have reported less support for abortion. Nigeria and Kenya both have a patriarchal tradition which favours male dominance. Women and adolescent girls constitute the most susceptible demography, facing heightened vulnerability, particularly in terms of poverty within households and communities (29). This vulnerability is further compounded by the detrimental cultural perceptions, and beliefs regarding gender roles, norms, and the empowerment of females. This has continued to fuel gender-based violence, lack of autonomy for women as well as poor reproductive health indices including low contraceptive use (29). It is not uncommon for adolescents growing in this environment to be socialised into such norms (29). Our study found a stronger endorsement of gender norms among VYAs in Nigeria relative to Kenya. While studies have found that children learn gender roles and norms from very young ages (29), several other factors including media can influence the adoption of these norms.

Our study revealed that endorsement of gender roles was significant with a positive attitude towards abortion. It is also important to note that the influence of gender roles/norms on abortion attitudes have been age-long with varied results. While there is a dearth of data on research of this nature in Africa, similar studies (31,32) from the Western world reveal contrary findings where an inverse relationship between the approval of traditional gender roles and support for abortion were observed. It is important to note however that none of these studies were conducted among VYA. Also, the gender roles considered in their studies were related more with women participation in economic / political activities. Conversely, (33) found gender norms to be a strong predictor of positive attitude towards abortion, while (34) on the other hand observed that gender norms and attitudes toward abortion are only loosely intertwined. No doubt, there exists a relationship between gender norms and abortion attitudes. However, as posited by (35), a more intricate analysis of the specific gender-role attitudes influencing perceptions of women’s roles and rights within society is necessary to better comprehend how these attitudes relate to the opposition or endorsement of abortion.

Research on has suggested that adolescents’ attitudes towards abortion are influenced by a combination of factors, including gender norms, age, marital status, religion, cultural context, access to information and healthcare services as well as reasons for procuring abortion - people are more likely to have a positive attitude to traumatic abortion rather than elective abortion (36). As with any societal issue, it is important to consider the diversity of perspectives and experiences among adolescents when examining attitudes towards abortion.

## Conclusion

VYAs are at a critical developmental stage where gender norms and attitudes are learnt and internalized. Gender norms and attitudes play an important role in decision-making processes towards access to sexual and reproductive health services, including abortion services. At an age when behaviors and attitudes are being learnt, it is important for interventions addressing gender and SRH components to inculcate positive gender norms about SRH including abortion. Such interventions should begin at an early age before gender norms and beliefs are turned into practice or behaviour.

## Data Availability

All data produced are available online at: https://www.icpsr.umich.edu/web/ICPSR/studies/38392.

## Acknowledgement

We would like to acknowledge the Academy for Health Development (AHEAD) and APHRC for receiving a grant from the International Development Research Centre (IDRC) to conduct the study, the results of which are presented in this article. The data utilized in this research is stored in the ICPSR repository. We appreciate both organizations’ contribution to the study and the opportunity to use the data for this publication.

## Notes

**Conflict of Interest:** The Authors declare none.

### Competing Interest Statement

The authors have declared no competing interest.

### Funding Statement

The study was funded by the International Development Research Center (Canada) (108676-004).

### Author Declarations

Ethical Review Committee of the Ministry of Health of Osun State and Kenya

## REFERENCES

1. WHO. Abortion: Key Facts. https://www.who.int/news-room/fact-sheets/detail/abortion.[Internet]. 2021. xAvailable from: https://www.who.int/news-room/fact-sheets/detail/abortion

2. Ganatra B, Tunçalp Ö, Johnston HB, Johnson Jr BR, Gülmezoglu AM, Temmerman M. From concept to measurement: operationalizing WHO’s definition of unsafe abortion. Bull World Health Organ. 2014 Mar 1;92(3):155–155.

3. Espinoza C, Samandari G, Andersen K. Abortion knowledge, attitudes and experiences among adolescent girls: a review of the literature. Sexual and Reproductive Health Matters. 2020 Jan 1;28(1):1744225.

4. Atuhaire S. Abortion among adolescents in Africa: A review of practices, consequences, and control strategies. Int J Health Plann Mgmt [Internet]. 2019 Oct [cited 2023 Apr 30];34(4). Available from: https://onlinelibrary.wiley.com/doi/10.1002/hpm.2842

5. Kabagenyi A, Jennings L, Reid A, Nalwadda G, Ntozi J, Atuyambe L. Barriers to male involvement in contraceptive uptake and reproductive health services: a qualitative study of men and women’s perceptions in two rural districts in Uganda. Reprod Health. 2014 Dec;11(1):21.

6. Larsson S, Eliasson M, Klingberg Allvin M, Faxelid E, Atuyambe L, Fritzell S. The discourses on induced abortion in Ugandan daily newspapers: a discourse analysis. Reprod Health. 2015 Dec;12(1):58.

7. Bell SO, Omoluabi E, OlaOlorun F, Shankar M, Moreau C. Inequities in the incidence and safety of abortion in Nigeria. BMJ Glob Health. 2020 Jan;5(1):e001814.

8. Ushie BA, Juma K, Kimemia G, Ouedraogo R, Bangha M, Mutua M. Community perception of abortion, women who abort and abortifacients in Kisumu and Nairobi counties, Kenya. Bourgeois D, editor. PLoS ONE. 2019 Dec 12;14(12):e0226120.

9. Abiola AH, Oke OA, Balagun MR, Olatona FA, Adegbesan-Omilabu MA. Knowledge, attitude, and practice of abortion among female students of two public senior secondary schools in Lagos Mainland Local Government Area, Lagos. J Clin Sci. 2016;13:82–7.

10. Rogers AA, Nielson MG, Santos CE. Manning up while growing up: A developmental-contextual perspective on masculine gender-role socialization in adolescence. Psychology of Men & Masculinities. 2021 Apr;22(2):354–64.

11. Bian L, Leslie SJ, Cimpian A. Gender stereotypes about intellectual ability emerge early and influence children’s interests. Science. 2017 Jan 27;355(6323):389–91.

12. Kollmayer M, Schober B, Spiel C. Gender stereotypes in education: Development, consequences, and interventions. European Journal of Developmental Psychology. 2018 Jul 4;15(4):361–77.

13. Weitzman L. “Sex-Role Socialization.” In Jo Freeman (Ed.) Women: A Feminist. 1975.

14. Mosley EA, Schulz AJ, Harris LH, Anderson BA. South African abortion attitudes from 2007-2016: the roles of religiosity and attitudes toward sexuality and gender equality. Women & Health. 2020 Aug 8;60(7):806–20.

15. Yorulmaz Ö. Are the Determinants of Attitudes Toward Abortion Changing in Turkiye? Sex Res Soc Policy [Internet]. 2023 Mar 15 [cited 2023 Aug 18]; Available from: https://link.springer.com/10.1007/s13178-023-00806-2

16. Lizotte MK. The Abortion Attitudes Paradox: Model Specification and Gender Differences. Journal of Women, Politics & Policy. 2015 Jan 2;36(1):22–42.

17. Akinlusi FM, Rabiu KA, Olawepo TA, Adewunmi AA, Ottun TA, Akinola OI. Sexual assault in Lagos, Nigeria: a five year retrospective review. BMC Women’s Health. 2014 Dec;14(1):115.

18. Ramathuba DU, Ngambi D, Khoza LB, Ramakuela NJ. Knowledge, attitudes and practices regarding cervical cancer prevention at Thulamela Municipality of Vhembe District in Limpopo Province. African Journal of Primary Health Care and Family Medicine. 2016;8(2):1–7.

19. Bankole A, Biddlecom A, Singh S, Guiella G, Zulu E. Sexual behavior, knowledge and information sources of very young adolescents in four sub-Saharan African countries. African journal of reproductive health. 2017;11(3):28–43.

20. Maina BW, Orindi BO, Sikweyiya Y, Kabiru CW. Gender norms about romantic relationships and sexual experiences among very young male adolescents in Korogocho slum in Kenya. Int J Public Health. 2020 May;65(4):497–506.

21. Woog V, Kagesten A. The sexual and reproductive health needs of very young adolescents aged 10–14 in developing countries: what does the evidence show?. [Internet]. Guttmacher Institure; 2017 p. 52. Available from: https://www.guttmacher.org/report/srh-needs-very-young-adolescents-in-developing-countries

22. Cleeve A, Faxelid E, Nalwadda G, Klingberg-Allvin M. Abortion as agentive action: reproductive agency among young women seeking post-abortion care in Uganda. Culture, Health & Sexuality. 2017 Nov 2;19(11):1286–300.

23. Moreau C, Li M, De Meyer S, Vu Manh L, Guiella G, Acharya R, et al. Measuring gender norms about relationships in early adolescence: Results from the global early adolescent study. SSM - Population Health. 2019 Apr;7:100314.

24. Nattino G, Pennell ML, Lemeshow S. Assessing the goodness of fit of logistic regression models in large samples: A modification of the HosmerLLemeshow test. Biometrics. 2020 Jun;76(2):549–60.

25. Bello BM, Fatusi AO, Adepoju OE, Maina BW, Kabiru CW, Sommer M, et al. Adolescent and Parental Reactions to Puberty in Nigeria and Kenya: A Cross-Cultural and Intergenerational Comparison. Journal of Adolescent Health. 2017 Oct;61(4):S35–41.

26. Mmari K, Moreau C, Gibbs SE, De Meyer S, Michielsen K, Kabiru CW, et al. ‘Yeah, I’ve grown; I can’t go out anymore’: differences in perceived risks between girls and boys entering adolescence. Culture, Health & Sexuality. 2018 Jul 3;20(7):787–98.

27. Frederico M, Michielsen K, Arnaldo C, Decat P. Factors Influencing Abortion Decision-Making Processes among Young Women. IJERPH. 2018 Feb 13;15(2):329.

28. Makleff S, Wilkins R, Wachsmann H, Gupta D, Wachira M, Bunde W, et al. Exploring stigma and social norms in women’s abortion experiences and their expectations of care. Sexual and Reproductive Health Matters. 2019 Nov 29;27(3):50–64.

29. Mochache V, Wanje G, Nyagah L, Lakhani A, El-Busaidy H, Temmerman M, et al. Religious, socio-cultural norms and gender stereotypes influence uptake and utilization of maternal health services among the Digo community in Kwale, Kenya: a qualitative study. Reprod Health. 2020 Dec;17(1):71.

30. Adaji SE, Linnea U, Warennius AA, Ongany AA, Faxelid EA. The attitudes of Kenyan in-school adolescents toward sexual autonomy. Afr J Reprod Health. 2010;14(1):33–42.

31. Sahar G, Karasawa K. Is the Personal Always Political? A Cross-Cultural Analysis of Abortion Attitudes. Basic and Applied Social Psychology. 2005 Dec;27(4):285–96.

32. Wang G zhen, Buffalo MD. Social and cultural determinants of attitudes toward abortion: a test of Reiss’ hypotheses. The Social Science Journal. 2004 Mar 1;41(1):93–105.

33. Dugger K. Race differences in the determinants of support for legalized abortion. Social Science Quarterly. 1991;72(3570–587).

34. Granberg D, Granberge BW. Social bases of support and opposition to legalized abortion. Perspectives on abortion. 1985;191–204.

35. Osborne D, Huang Y, Overall NC, Sutton RM, Petterson A, Douglas KM, et al. Abortion Attitudes: An Overview of Demographic and Ideological Differences. Political Psychology. 2022 Nov;43(S1):29–76.

36. Newton SL, Hebert LE, Nguyen BT, Gilliam ML. Negotiating Masculinity in a Women’s Space: Findings from a Qualitative Study of Male Partners Accompanying Women at the Time of Abortion. Men and Masculinities. 2020 Apr;23(1):65–82.

